# Preconception metabolic-bariatric surgery and child health outcomes: Identification and cohort profile of the POSIT study protocol

**DOI:** 10.64898/2026.04.22.26351521

**Authors:** Jonathan Q. Purnell, Darios Getahun, Kimberly K. Vesco, Sijia Qiu, Jiaxiao M. Shi, Carmen P. Wong, Padma Koppolu, Theresa M. Im, Caryn E. Oshiro, Janne Boone-Heinonen

## Abstract

Preconception weight loss by metabolic-bariatric surgery (MBS) improves maternal-fetal outcomes, but little is known about its impact on offspring growth and health. The preconception bariatric surgery and child health outcomes (POSIT) study aims to estimate the effects of maternal MBS-induced preconception weight loss on infant and childhood body size, growth, and related outcomes. This report presents the methods used to construct the POSIT cohort and its baseline characteristics. This retrospective cohort study sampled members from a United States healthcare system aged 18 and older with a singleton, live birth to create three study groups: 1) a treatment group including women who underwent preconception MBS and subsequently became pregnant (n=1,374); 2) a control group matched to the MBS pre-surgery body mass index (BMI) (pre-surgery controls, n=13,740); and 3) a second control group matched to the MBS post-surgical, pre-pregnancy BMI (pre-pregnancy controls, n=13,740). MBS and pre-surgery BMI controls showed slight imbalances in that pre-surgery BMI controls were on average ∼6 months younger, had 0.6 lower BMI (44.5 kg/m^2^) at the time of their pregnancy and were more likely to have become pregnant in earlier years than the MBS group prior to surgery. MBS and pre-pregnancy controls had comparable age (mean ± SD 33 ± 5 years), pre-pregnancy BMI (33 ± 6 kg/m^2^), and year of delivery. Following matching, the MBS group had similar socioeconomic and health disparities as the pre-surgery control group, and both were worse than pre-pregnancy control group. Pregestational maternal comorbidity index improved after MBS and matched the pre-pregnancy controls. Upon extraction of offspring growth patterns and mediation analyses of maternal weight loss and metabolic responses to MBS, study findings will investigate effects of preconception weight loss by MBS on short- and long-term child health outcomes. Results will guide future studies focusing on improving maternal preconception weight and maternal-fetal outcomes.

## Introduction

During the past three decades, U.S. obesity prevalence has dramatically risen to over 40% of the general population by 2018,[1] a disproportionate number of whom identify as Black or Hispanic [2] and including an greater number of women entering pregnancy.[3] Animal and human studies suggest that maternal obesity increases an offspring’s risk for developing obesity and associated chronic diseases in adulthood.[4–10] However, disappointing results from recent studies of lifestyle interventions *during pregnancy* in women with obesity[11–14] have shifted interventional priorities to the *preconception* period.

Lifestyle approaches to reduce preconception weight result in modest average total body weight losses of 4% to 8%.[15, 16] Obesity medications can increase this weight loss to ∼6% to 20%,[17] but, because of inadequate safety data, they are not recommended for use in women attempting conception or during pregnancy.[18, 19] In contrast, total weight loss after metabolic-bariatric surgery (MBS) procedures averages 20% to 30%, is sustained long-term (7 years and longer), and significantly reduces obesity-related complications.[20–22] As a result, utilization of MBS procedures in women with severe obesity is rising, including those of childbearing age.[23]

Recent reports [24] and meta-analyses[25] of women who have undergone MBS have documented improved maternal metabolic health and neonatal outcomes in subsequent pregnancies. Notably, these reports also demonstrated increased risk for small for gestational age births, which may have opposing effects on the offspring’s longer-term obesity and chronic disease risk.[26] However, few studies have tracked weight trajectories of infants and children born to women who have undergone MBS, with several showing promising results but no clear conclusions of benefit or harm.[27–33] These studies were often limited by small sample sizes (ranging from 40 to 400), lacked control groups of weight-matched women, or did not include minoritized racial and ethnic groups. Thus, there is a critical need to understand the risks and benefits of preconception surgical weight loss on long-term child health outcomes, the results of which could affect weight management priorities and approaches for women with obesity in their child-bearing years.

The objectives of the PreconceptiOn bariatric Surgery and chIld health ouTcomes (POSIT) study are to estimate the effects of preconception MBS on offspring weight and weight-related health up to age 5, and to determine the extent to which pregnancy conditions mediate associations between MBS and these offspring outcomes. We will conduct these aims in a large, racially and ethnically diverse, cohort of women and their children derived from electronic health record (EHR) data from a multi-site integrated healthcare network. We will examine three study groups: (1) a treatment group of women who underwent preconception MBS and subsequently became pregnant, (2) a control group of women with no history of bariatric surgery who are matched to the MBS pre-surgical body mass index (BMI) (pre-surgery controls), and (3) a control group of women with no history of bariatric surgery matched to the MBS post-surgical, pre-pregnancy BMI (pre-pregnancy controls). Herein, we present the methods used to construct this cohort of three study groups of mother-child dyads and describe baseline characteristics of the cohort.

## Methods

### Study population

The POSIT study includes patients from Kaiser Permanente’s (KP) Hawaii, Northwest, and Southern California regions (KPHI, KPNW, KPSC). We used integrated administrative and EHR data from KP’s Virtual Data Warehouse (VDW) across the three KP regions. The VDW was created to facilitate multi-site research projects. Local variables derived from integrated EHR data with information from outpatient encounters, hospital admissions, laboratory results, prescriptions, and claims, are standardized using consistent naming, definitions, and formats. This research was reviewed and approved by the Oregon Health & Science University and Kaiser Permanente Integrated Institutional Review Boards with a waiver of informed consent. KPSC received approval from the California Health and Human Services Agency and California Department of Public Health Center for Health Statistics and Informatics for the use of state birth files.

To construct the POSIT cohort, we identified pregnancies from pregnancy episodes within Epic, which are initiated by a medical provider or their support staff at the onset of an individual’s prenatal care. Pregnancy episodes include variables indicating date of last menstrual period (LMP), estimated delivery date (EDD), pregnancy outcome, gestational age at delivery (when known), and other pregnancy-level information. Estimated pregnancy onset date (POD) is updated throughout pregnancy based on the best clinical estimates of gestational age including ultrasound. Data extraction across sites was performed by distributing a single SAS program to all KP sites; sites made minor modifications where needed to extract comprehensive data for preconception, pregnancy, delivery, postpartum, and child health; and limited data files were transferred to a single site for analysis.

A base population of eligible pregnancies was identified with an initial set of inclusion criteria: known pregnancy onset between 1/1/2006 and 12/31/2020, known pregnancy outcome date on or before 12/31/2020, and known pregnancy type (e.g. livebirth) without overlap with other pregnancy episodes; maternal age 18 years or older at pregnancy onset; at least one valid height and pre-pregnancy weight; and a singleton live birth with a linked child record in the EHR (**Fig 1**). The surgical and control groups were selected from the base population described below (Study groups selection).

**Fig 1.**
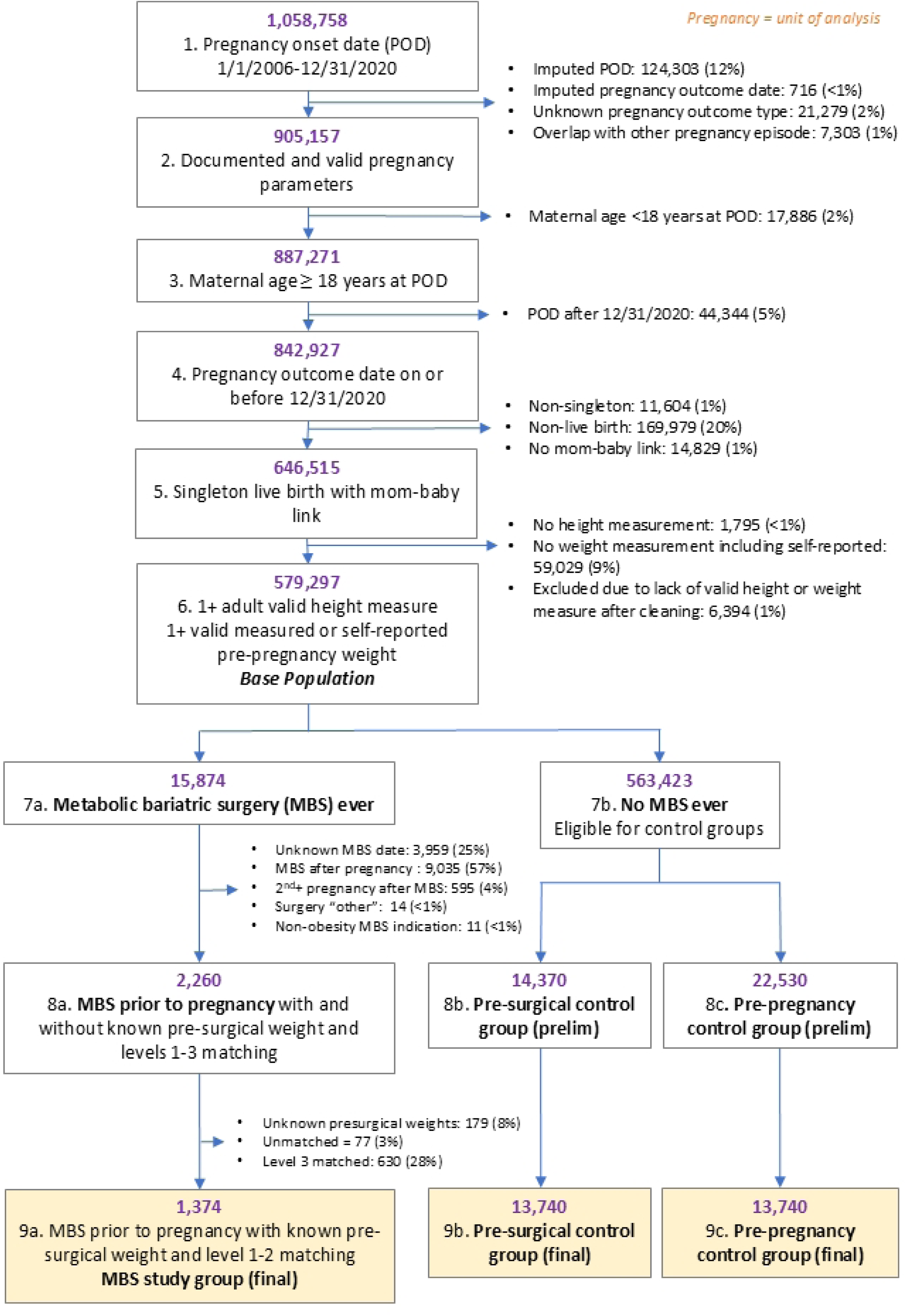
Flow diagram describing the identification of study groups. Matching criteria for steps 8 and 9 included maternal age, body mass index (BMI), calendar year of pregnancy onset, and KP site. Only women who underwent Roux-en-Y gastric bypass or sleeve gastrectomy were eligible; “other” surgeries included laparoscopic gastric banding and biliopancreatic diversion/duodenal switch. Phase 1 matching for both pre-surgery controls (PSC) and pre-pregnancy controls (prepregnancy control): match pair ratio was set to 1:10 with the following caliper widths: ±2 years of age, ±2 calendar years, ±2 BMI units. In phase 2 matching, bariatric surgery group members without at least 1 matched control were matched using larger caliper widths: ±5 years of age, ±5 calendar years, ± 4 BMI units. Maternal age and pregnancy outcome year were weighted at 1; BMI was weighted at 2; maximum allowable distance for all the 3 variables was ±2. Level 2 matching for unmatched PSC only: match ratio set to 1:1; Level 3 matching occurred only for unmatched PSC with a match ratio set to 1:1 and BMI matching calipers extended to ±10 BMI units.

### Data Sources

Pregnancies in the base population were linked to clinical data for the child and state birth certificate data. In the KP VDW, pregnancies are routinely linked to child clinical data using a validated algorithm including maternal medical record number, infant birth date, and pregnancy outcome.[34] Within the POSIT base population, 99% of live births were linked to child clinical records. State birth certificate data were obtained by each state (KPNW: OR; KPSC: CA; KPHI: HI). Each site conducted probabilistic linkage to the POSIT study populations [35] and constructed relevant study variables synchronized across states and years. Patient residential addresses are continuously updated and geocoded in the VDW, mapped to geographic units (e.g., county, census tract) linked to US Census and other national data sources.[36]

### Study variables

#### Maternal anthropometry

All clinically recorded adult heights and maternal weights from 180 days prior to POD through 84 days after POD were extracted from clinical encounter data through 5/30/2024. Same-day measures were aggregated by averaging if they differed by less than 10%. The data were then cleaned using the validated growthcleanr package in R (adult module)[37] including multiple same day measures with greater than 10% of difference; 1.5% of all height and 0.7% of weight measures flagged as implausible and excluded. ***Maternal height*** was calculated as the median of all post-cleaned height measures recorded from 18 years of age through the data extraction date. ***Maternal pre-pregnancy weight*** was selected as the closest weight prior to the POD among cleaned weight measures from 180 days prior to the POD. If no valid weight was recorded before POD, patient-reported pre-gravid weight recorded in the pregnancy episode table was used (n=XX, XX%); pre-gravid weight is highly correlated with measured pre-pregnancy weight (r = 0.99).[38] If both pre-POD weights and pre-gravid weight records were missing, the closest weight after POD within 0 to 84 days window was selected (n=XX, XX%).[38] ***Maternal pre-pregnancy BMI*** was calculated from pre-pregnancy weight in kilograms (kg) and height in meters squared (m^2^).

#### Metabolic-bariatric surgery (MBS)

Metabolic bariatric surgery events were identified based on the International Classification of Diseases (ICD-9) and (ICD-10) Revisions diagnosis codes and Current Procedural Terminology (CPT-4) procedure codes recorded during clinical encounters (**S1 and S2 Tables**), including all historical data for patients in the base population. The date associated with the procedure code was used as the surgery date. ***Pre-surgery BMI*** *was* calculated in those who had a recorded MBS using height and the highest non-pregnancy weight within 12 months prior to the surgical date. This timeframe, rather than the recorded weight at the time of surgery, was selected because it acknowledges that some surgical programs or health insurance companies mandate pre-surgery weight loss prior to procedure eligibility or coverage approval, typically within six months of the MBS procedure. Any weights recorded during a pregnancy were excluded from this determination. Procedure-related variables included MBS type (Roux-en-Y gastric bypass or sleeve gastrectomy), time from surgery date to pregnancy onset date, and weight change from the maximum presurgical weight to the weight at the pregnancy onset date.

#### Maternal Demographic and Baseline Health Characteristics

Maternal age at delivery, calendar year of birth, insurance type at time of delivery (Medicaid, Medicare, commercial, other) and prenatal care initiation (trimester 1, 2, 3, or none/unknown) were determined from the VDW. Tobacco use (smoker, nonsmoker, missing) was determined primarily from data collected at clinical encounters: preconception smoking was defined as ≥1 report of tobacco use, non-smoking as 0 reports of tobacco use and ≥1 report of no tobacco use, otherwise missing. If tobacco use was missing based on VDW data, birth certificate data were used as a secondary source. Race & ethnicity (Hispanic, non-Hispanic, Black or African American, Asian, American Indian or Alaska Native, Native Hawaiian or Pacific Islander, unknown) and parity (nulliparous, multiparous, unknown) were determined from the birth certificate, or, if not available, the VDW. Neighborhood median household income, neighborhood household percent with income below poverty, and neighborhood deprivation index [39] were extracted from the VDW for the residential location with the longest duration in the 720 days (2 years) prior to POD. Neighborhood deprivation index (NDI) was used to indicate neighborhood-level socioeconomic status (SES), defined based on the census tract of residence spanning the longest duration in the preconception time window. If the participant resided at 2 addresses for same duration, we selected the location recorded closest to POD. We used Census data from the year of pregnancy onset, except for 2006-2011, for which we used the earliest NDI data available in the VDW (2012).

The maternal comorbidity index [40] is a validated [41] score comprising 20 maternal conditions, including pregestational hypertension and diabetes, and maternal age that summarizes the burden of maternal illness during pregnancy through 30 days postpartum. In the present study this index was modified to encompass the three months before pregnancy through the end of the first trimester and excluded pregnancy-related conditions. Pregestational hypertension was determined using ICD 9/10 codes starting 1 year prior to pregnancy onset date up through 20 weeks gestation, with at least one inpatient diagnosis or at least two emergency department/outpatient diagnoses.

### Study groups selection

#### Selection of the MBS group

Among patients with eligible pregnancies represented in the base population (**Fig 1**), those with a record of MBS were identified (n=15,874). Of these, 3,959 (25%) without a known surgical date were excluded. The first (initial) observed pregnancy after MBS for each woman was chosen as the index pregnancy. We examined only the index pregnancy for women in the MBS group; 9,035 pregnancies (57%) that occurred prior to MBS and 595 (4%) non-initial pregnancies after MBS were not examined in this study. Only women with Roux-en-Y gastric bypass (RYGB) and laparoscopic sleeve gastrectomy (LSG) procedures for MBS indication were included, excluding 14 (<1%) women who underwent laparoscopic gastric banding or biliopancreatic diversion/duodenal switch (listed as Surgery “other”) and 11 (<1%) with surgeries conducted for non-MBS indication (e.g., RYGB for severe esophageal reflux without obesity). Following these exclusions, 2,260 eligible women were selected for the MBS group.

#### Selection of the matched control groups

Among the remaining base population of eligible pregnancies after identification of the MBS group, two matched control groups were selected using the following four criteria: KP site, calendar year of pregnancy onset, maternal age, and either pre-surgery or pre-pregnancy BMI (**Figs 1 and 2**). The ***Pre-surgery Control Group*** consists of women with no history of MBS whose pre-pregnancy BMI was used to match with the pre-surgery BMI of women in the MBS group. This group was chosen to approximate expected infant and child weight outcomes of women with severe obesity if they had not undergone MBS-induced weight loss (**Fig 2**). The ***Pre-pregnancy Control Group*** consists of women with no history of MBS whose pre-pregnancy BMI was used to match with the post-surgical, pre-pregnancy BMI of women in the MBS group. This group was chosen to approximate infant and child weight outcomes expected after their women attain a new, lower weight (**Fig 2**). Pre-surgery and pre-pregnancy control groups were selected in two independent matching processes; that is, a given pregnancy could be assigned into both control groups. If a participant was selected for the MBS group, their pre-MBS pregnancies were not eligible for selection into the control group.

**Fig 2.**
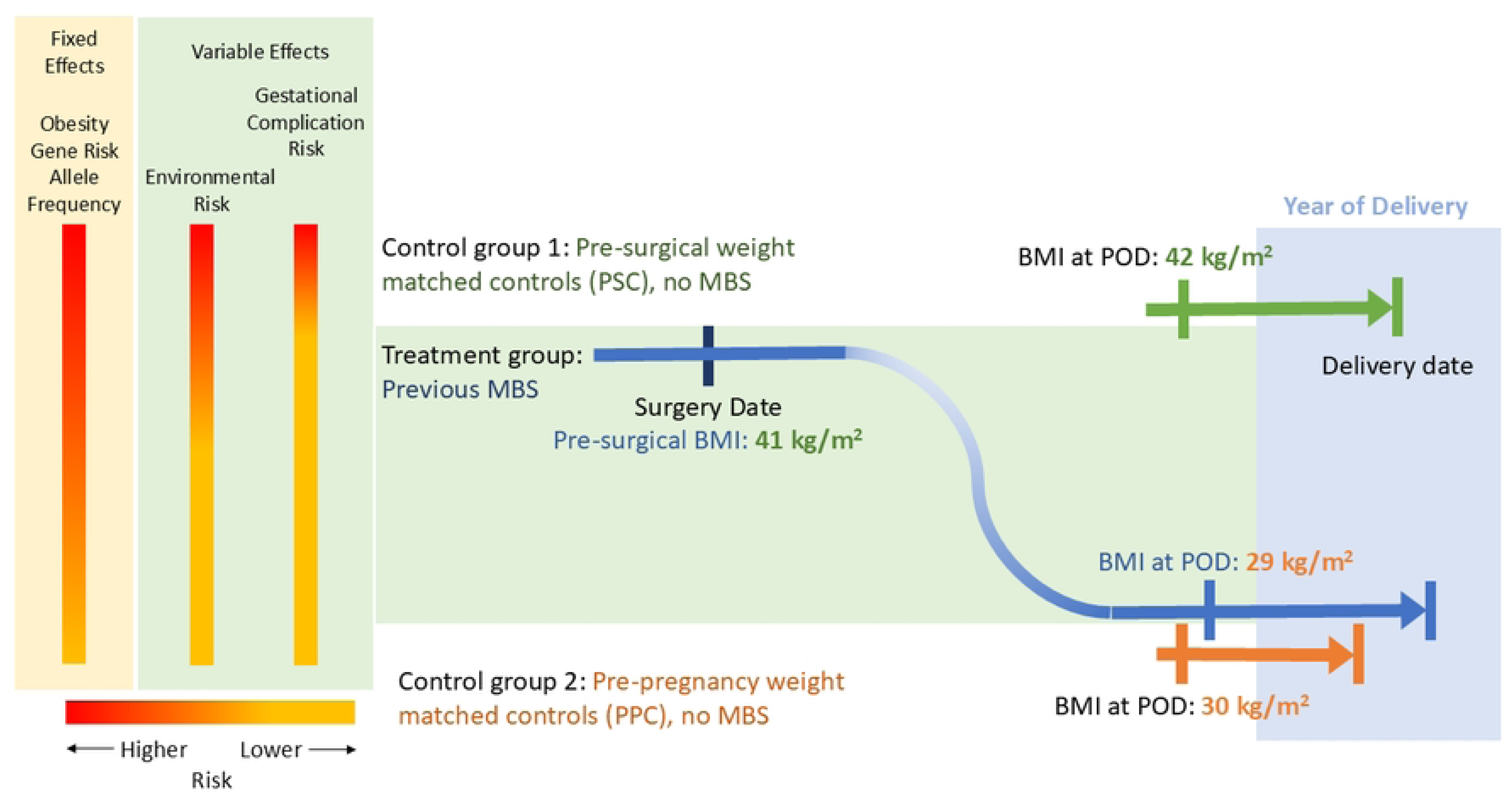
Matching example of control groups to the metabolic-bariatric surgery (MBS) group. Matching criteria included maternal age, body mass index (BMI) according to either pre-surgery or pre-pregnancy weights of the MBS group, year of delivery, and Kaiser Permanente study site. The highest obesity genetic allele burden (fixed effect) is predicted to occur in the MBS and PSC cohorts. Environmental influences may vary, but differences between groups are hoped to be minimized by matching to Kaiser site and year of pregnancy with gestational complication risks for maternal glycemia and hypertensive disorders of pregnancy being highest in the PSC group compared to MBS and PPC cohorts. POD: pregnancy onset date. PSC: pre-surgical controls. PPC: pre-pregnancy controls.

After excluding 179 women in the MBS group who did not have identifiable pre-surgery weights, we used weighted, nearest-neighbor matching with unequal weighting for age, year, and BMI (1:1:2 weight, respectively) and without replacement, aiming for a 1:10 MBS to control ratio. In phase 1, caliper ranges of ±2 years of age, ±2 calendar years, and ±2 BMI units were used in the matching process. However, due to very high pre-surgery BMI values observed in the MBS group, matched pre-surgery controls were not identified for all MBS cases using our primary matching criteria. In phase 1, a total of 1181 (52%) MBS cases were matched with 10 pre-surgery controls; 1002 (44%) were matched with 1-9 pre-surgery controls. Therefore, we implemented 2 additional matching phases to minimize the exclusion of those with extreme pre-surgery BMI values from analyses. In phase 2, wider caliper ranges were applied for MBS cases not achieving a 1:10 full match ratio in phase 1: ±5 years of age, ±5 calendar years, ±4 BMI units. In phase 2, 193 MBS cases (8.5%) were matched to 10 full controls. When combined with phase 1 matched controls, this resulted in 1,374 MBS cases and 13,740 pre-surgery controls (**Fig 1**, **Table 1**). Phase 3 matching used a 1:1 MBS to control the matching ratio with less restrictive criteria: BMI within ±10 units and KP site but omitting maternal age and calendar year of POD criteria. Phase 3 matching resulted in an additional 630 MBS cases (27.9%) matching pre-surgical controls (**S3 Table**).

**Table 1.** Baseline and matching characteristics of final POSIT cohorts.

| Socio-demographic and medical characteristics | Metabolic-Bariatric Surgery Group | Pre-surgery BMI Control Group | *P*-value | ASD<sup>1</sup> | Pre-pregnancy BMI Control Group | *P*-value | ASD<sup>1</sup> |
| --- | --- | --- | --- | --- | --- | --- | --- |
|  | N=1,374 | N=13,740 | MBS vs PSC | MBS vs PSC | N=13,740 | MBS vs PPC | MBS vs PPC |
| **Maternal age, years, Mean (SD)** | 32.65 (5.07) | 32.07 (5.10) | <0.01 | 0.12 | 32.6 (5.04) | 0.93 | <0.01 |
| **Maternal age, years, n (%)** |  |  | <0.01 | 0.10 |  | 0.99 | <0.01 |
| 18-25.9 | 134 (9.8) | 1,576 (11.5) |  |  | 1,355 (9.9) |  |  |
| 26-34.9 | 716 (52.1) | 7,546 (54.9) |  |  | 7,155 (52.1) |  |  |
| ≥35 | 524 (38.1) | 4,618 (33.6) |  |  | 5,230 (38.1) |  |  |
| **Pre-surgical BMI and matched BMI<sup>2</sup>, Mean (SD)** | 45.08 (4.91) | 44.51 (4.84) | <0.01 | 0.12 | -- | -- | -- |
| **Pre-surgical BMI and matched BMI<sup>2</sup>, n (%)** |  |  |  |  |  |  |  |
| 30-34.9 | 10 (0.7) | 100 (0.7) | 0.96 | <0.01 | -- |  |  |
| 35-39.9 | 113 (8.2) | 1,160 (8.4) |  |  | -- |  |  |
| ≥40 | 1,251 (91.0) | 12,480 (90.8) |  |  | -- |  |  |
| **Pre-pregnancy BMI and matched BMI<sup>2</sup>, kg/m<sup>2</sup>, Mean (SD)** | 33.02 (5.94) |  |  |  | 33.02 (5.93) | 0.99 | <0.01 |
| **Pre-pregnancy BMI and matched BMI<sup>2</sup>, kg/m<sup>2</sup>, n (%)** |  |  |  |  |  |  |  |
| 18.5-24.9 | 58 (4.2) |  |  |  | 580 (4.2%) | 1.00 | <0.01 |
| 25-29.9 | 356 (25.9) |  |  |  | 3,561 (25.9) |  |  |
| 30-34.9 | 475 (34.6) |  |  |  | 4,750 (34.6) |  |  |
| 35-39.9 | 307 (22.3) |  |  |  | 3,069 (22.3) |  |  |
| ≥40 | 178 (13) |  |  |  | 1,780 (13.0) |  |  |
| **KP sites, n (%)** |  |  | 1.00 | 0.00 |  | 1.00 | 0.00 |
| Hawaii | 26 (1.9) | 260 (1.9) |  |  | 260 (1.9) |  |  |
| Northwest | 47 (3.4) | 470 (3.4) |  |  | 470 (3.4) |  |  |
| Southern California | 1,301 (94.7) | 13,010 (94.7) |  |  | 13,010 (94.7) |  |  |
| **Calendar year of POD, n (%)** |  |  | <0.01 | 0.27 |  | 0.91 | 0.01 |
| 2006 - 2009 | 49 (3.6) | 1316 (9.6) |  |  | 518 (3.8) |  |  |
| 2010 - 2015 | 655 (47.7) | 5,437 (39.6) |  |  | 6,499 (47.3) |  |  |
| 2016 - 2020 | 670 (48.8) | 6,987 (50.9) |  |  | 6,723 (48.9) |  |  |

Comparisons include the treatment group who underwent pre-conception metabolic-bariatric surgery, a pre-surgery control group matched to pre-MBS BMI, and a pre-pregnancy control group matched to BMI at the time of pregnancy.
Values for comparison analysis on matching body mass index's (BMI's) between MBS and pre-surgery control group shown in shaded light grey.
Abbreviations: KP: Kaiser Permanente; MBS: Metabolic-Bariatric Surgery; NH: Non-Hispanic; POD: Pregnancy Onset Date; POSIT: Preconception Bariatric Surgery and Child Health Outcomes; PPC: Pre-Pregnancy Control; PSC: Pre-Surgical Control; SD: Standard Deviation.
Groups were matched on BMI, KP site, age at delivery, and delivery year.
<sup>1</sup> Absolute standardized difference (ASD) was calculated to assess the balance of covariates. Covariates with ASD ≤0.15 implies good balance.
<sup>2</sup> Defined as highest BMI within 12 months prior to the surgical date for MBS group; as closest pre-pregnancy BMI from 180 days prior to pregnancy onset date for control groups.

Only cases (n=1,374) and their controls matched (n=13,740 each group) in phases 1 and 2 were included in the final analysis (**Fig 1**, **Table 1**). For sensitivity analysis, we included MBS cases with full and partial matching from matching phases 1 to 3 (**Fig 1, S3 and S4 Tables**). Larger, more inclusive, pre-surgery control and pre-pregnancy control groups were selected independently

#### Statistical Analysis

Descriptions of maternal group characteristics include continuous covariates summarized by mean and standard deviation and compared using Wilcoxon rank-sum test. Categorical variables are presented as absolute numbers and percentages with *P*-values for the χ2 test or Fisher’s exact test, as appropriate. Absolute Standardized Difference (ASD), calculated as the absolute difference divided by the pooled standard deviation, was used to evaluate the covariate balance among the matched groups.

## Results

### Maternal Baseline Matching Variables

The primary cohort (**Table 1**) included 1,374 MBS women matched to 13,740 pre-surgical controls and 13,740 pre-pregnancy controls. In general, balance was achieved between the study groups for the matching variables (age, BMI, calendar year of delivery, and KP site), indicated by ASD <0.15. The mean ages of the MBS, pre-surgery control, and pre-pregnancy control groups were all within 1 year of each other. The mean pre-surgery BMI for the MBS group (45.08 kg/m^2^) was within 1 BMI unit of the pre-surgery control group (see shaded areas for comparison in **Table 1**) (44.51 kg/m^2^) and the MBS group pre-pregnancy BMI (33.02 kg/m^2^) was identical to the pre-pregnancy control group (33.02 kg/m^2^). Also identical were the balance of recruitment between KP sites and the calendar year of POD between the MBS and pre-pregnancy group. Larger differences in calendar year between the MBS group and pre-surgery control group reflected a broader search including earlier calendar years of POD needed to identify the pre-surgery control group (ASD 0.27). (**Table 1**).

Similar results for matching variables were found between groups identified in the phase 2/3 matching (**S3 Table**), but the imbalances were greater, especially for maternal BMI comparing MBS (48.38 kg/m^2^) versus the pre-surgery control group (44.53 kg/m^2^) due to the skewing of pre-surgical BMI’s towards very high levels in the MBS group.

#### Maternal Baseline Demographic and Health Characteristics

The most common MBS procedure was sleeve gastrectomy (62.4%) with the remainder undergoing gastric bypass (37.6%) (**Table 2**). The mean ± SD interval time from surgery to pregnancy was 889 ± 698 days, and the mean ± SD percent weight change between pre-surgery and pre-pregnancy was -26.6% ± 11.6%.

**Table 2.** Baseline demographic and maternal health characteristics of final POSIT cohorts.

| Socio-demographic and medical characteristics | Metabolic-Bariatric Surgery Group | Pre-surgery Control Group | *P*-value | ASD<sup>1</sup> | Pre-pregnancy Control Group | *P*-value | ASD<sup>1</sup> |
| --- | --- | --- | --- | --- | --- | --- | --- |
|  | N=1374 | N=13740 | MBS vs PSC | MBS vs PSC | N=13740 | MBS vs PPC | MBS vs PPC |
| **MBS Surgical Type, n (%)** |  |  |  |  |  |  |  |
| Roux-en-Y Gastric Bypass | 516 (37.6%) | -- | -- | -- | -- | -- | -- |
| Sleeve Gastrectomy | 858 (62.4%) | -- | -- | -- | -- | -- | -- |
| **Race and Ethnicity, n (%)** |  |  | <0.01 | 0.14 |  | <0.01 | 0.40 |
| NH White | 293 (21.3) | 3031 (22.1) |  |  | 3271 (23.8) |  |  |
| NH Black or African American | 222 (16.2) | 1783 (13.0) |  |  | 1163 (8.5) |  |  |
| NH Asian | 13 (0.9) | 328 (2.4) |  |  | 1057 (7.7) |  |  |
| Hispanic | 792 (57.6) | 7982 (58.1) |  |  | 7679 (55.9) |  |  |
| Other / Unknown | 54 (3.9) | 616 (4.5) |  |  | 570 (4.1) |  |  |
| **Insurance Type, n (%)** |  |  |  |  |  |  |  |
| Medicaid<sup>2</sup> | 358 (26.1) | 2,937 (21.4) | <0.01 | 0.11 | 2,231 (16.2) | <0.01 | 0.24 |
| Medicare<sup>2</sup> | 17 (1.2) | 66 (0.5) | <0.01 | 0.08 | 25 (0.2) | <0.01 | 0.13 |
| Commercial insurance<sup>2</sup> | 1,000 (72.8) | 10,495 (76.4) | <0.01 | 0.08 | 10,952 (79.7) | <0.01 | 0.16 |
| Other insurance<sup>2</sup> | 736 (53.6) | 7,212 (52.5) | 0.45 | 0.02 | 6,927 (50.4) | 0.03 | 0.06 |
| **Neighborhood median household income, USD, Mean (SD)** | \$59,083 (21,749) | \$60,263 (22,704) | 0.14 | 0.01 | \$64,672 (24,877) | <0.01 | 0.24 |
| **Neighborhood median household income, USD, n (%)** |  |  | <0.01 | 0.29 |  | <0.01 | 0.37 |
| <\$40,000 | 254 (18.5) | 2,394 (17.4) | | | 1,984 (14.4) | | |
| \$40,000-\$59,999 | 551 (40.1) | 5,035 (36.6) | | | 4,412 (32.1) | | |
| \$60,000-\$79,999 | 337 (24.5) | 3,306 (24.1) | | | 3,549 (25.8) | | |
| \$80,000+ | 232 (16.9) | 2,460 (17.9) | | | 3,221 (23.4) | | |
| Unknown | 0 (0.0) | 545 (4.0) |  |  | 574 (4.2) |  |  |
| **Household Poverty, Mean (SD)** | 0.15 (0.10) | 0.15 (0.10) | 0.10 | 0.03 | 0.13 (0.10) | <0.01 | 0.19 |
| **NDI, Mean (SD)** | 0.70 (0.95) | 0.65 (0.98) | 0.01 | 0.06 | 0.45 (0.97) | <0.01 | 0.26 |
| **NDI Quartile<sup>3</sup>, n (%)** |  |  | <0.01 | 0.15 |  | <0.01 | 0.33 |
| Quartile 1 | 258 (18.8) | 3,376 (24.6) | <0.01 | 0.15 | 4,428 (32.2) | <0.01 | 0.33 |
| Quartile 2 | 334 (24.3) | 3,309 (24.1) |  |  | 3,341 (24.3) |  |  |
| Quartile 3 | 393 (28.6) | 3,487 (25.4) |  |  | 3,055 (22.2) |  |  |
| Quartile 4, most deprived | 389 (28.3) | 3,568 (26.0) |  |  | 2,916 (21.2) |  |  |
| **Timing of first prenatal care visits, n (%)** |  |  | <0.01 | 0.27 |  | <0.01 | 0.19 |
| 1st trimester | 1,263 (91.9) | 11,479 (83.5) |  |  | 11,886 (86.5) |  |  |
| 2nd trimester | 92 (6.7) | 1,657 (12.1) |  |  | 1,476 (10.7) |  |  |
| 3rd trimester | 8 (0.6) | 365 (2.7) |  |  | 270 (2.0) |  |  |
| Unknown or no prenatal care | 11 (0.8) | 239 (1.7) |  |  | 108 (0.8) |  |  |
| **Parity, Mean (SD)** | 0.99 (1.13) | 1.22 (1.24) | <0.01 | 0.19 | 1.21 (1.22) | <0.01 | 0.19 |
| **Parity, n (%)** |  |  | <0.01 | 0.21 |  | <0.01 | 0.22 |
| 0 | 589 (42.9) | 4,505 (32.8) |  |  | 4,461 (32.5) |  |  |
| ≥1 | 773 (56.3) | 9,143 (66.5) |  |  | 9,212 (67.0) |  |  |
| Unknown | 12 (0.9) | 92 (0.7) |  |  | 67 (0.5) |  |  |
| **Time from surgery to pregnancy onset date, days, Mean (SD)** | 889 (698) | -- | -- | -- | -- | -- | -- |
| **Post-surgery % Weight Change, lbs, Mean (SD)** | -26.6 (11.6) | -- | -- | -- | -- | -- | -- |
| **Tobacco use during pregnancy, n (%)** |  |  | 0.01 | 0.09 |  | 0.42 | 0.04 |
| Ever | 32 (2.3) | 534 (3.9) |  |  | 380 (2.8) |  |  |
| Never | 1,319 (96.0) | 12,996 (94.6) |  |  | 13,173 (95.9) |  |  |
| Unknown | 23 (1.7) | 210 (1.5) |  |  | 187 (1.4) |  |  |
| **Alcohol use,<sup>4</sup> n (%)** |  |  | <0.01 | 0.57 |  | <0.01 | 0.51 |
| Ever | 516 (37.6) | 3,823 (27.8) |  |  | 4,272 (31.1) |  |  |
| Never | 795 (57.9) | 6,686 (48.7) |  |  | 6,554 (47.7) |  |  |
| Unknown | 63 (4.6%) | 3,231 (23.5) |  |  | 2,914 (21.2) |  |  |
| **Pre-gestational Hypertension<sup>5</sup>, n (%)** |  |  | 0.99 | <0.01 |  | <0.01 | 0.22 |
| Yes | 142 (10.3) | 1,419 (10.3) |  |  | 624 (4.5) |  |  |
| No | 1,232 (89.7) | 12,321 (89.7) |  |  | 13,116 (95.5) |  |  |
| **Maternal Comorbidity Index<sup>6</sup>, Mean (SD)** | 0.47 (0.73) | 0.40 (0.65) | <0.01 | 0.11 | 0.44 (0.64) | 0.40 | 0.05 |
| **Maternal Comorbidity Index<sup>6</sup> Group, n (%)** |  |  | <0.01 | 0.11 |  | <0.01 | 0.09 |
| 0 | 867 (63.1) | 9,264 (67.4) |  |  | 8,758 (63.7) |  |  |
| 1 | 397 (28.9) | 3,681 (26.8) |  |  | 4,082 (29.7) |  |  |
| 2 | 92 (6.7) | 664 (4.8) |  |  | 818 (6.0) |  |  |
| 3 | 11 (0.8) | 97 (0.7) |  |  | 70 (0.5) |  |  |
| ≥4 | 7 (0.5) | 34 (0.2) |  |  | 12 (0.1) |  |  |

Comparisons include the treatment group who underwent pre-conception metabolic-bariatric surgery, a pre-surgery control group matched to pre-MBS BMI, and a pre-pregnancy control group matched to BMI at the time of pregnancy.
Abbreviations: MBS: Metabolic-Bariatric Surgery; NDI: Neighborhood Deprivation Index; NH: Non-Hispanic; PPC: Pre-Pregnancy Control; PSC: Pre-Surgical Control; SD: Standard Deviation; USD: US Dollar.
<sup>1</sup>Absolute standardized difference (ASD) was calculated to assess the balance of covariates. Covariates with ASD ≤0.15 implies good balance.
<sup>2</sup>Defined at the time of delivery.
<sup>3</sup>Possible range: -2 to 6.
<sup>4</sup>Defined in the two years prior to the pregnancy onset date.
<sup>5</sup>Defined as having at least one inpatient chronic hypertension diagnosis or two emergency department/urgent care diagnoses 1 year prior to pregnancy onset date to before 20 weeks into pregnancy.
<sup>6</sup>Defined 3 months before pregnancy through the end of 1st trimester excluding pregnancy-related conditions, modified from original definition (see text for explanation).[40, 41]

Race and ethnicity showed some group differences, with the MBS group including a greater percentage who self-identified non-Hispanic Black (16.2%) and fewer who identified as non-Hispanic White (21.3%) (ASD 0.14 and 0.40 compared to the pre-surgery and pre-pregnancy control groups, respectively) (**Table 2**). The highest percentage of women who identified as Asian was found in the pre-pregnancy control group (7.7% compared to 2.4% in the pre-surgery group and 0.9% in the MBS group). Insurance type, neighborhood median income, household poverty, and neighborhood deprivation indices were similar for the MBS versus pre-surgery groups. In contrast, the pre-pregnancy control group had fewer enrolled in Medicaid and more in commercial insurance and reported higher (on average) household income and lower household poverty and neighborhood deprivation indices than both the MBS and pre-surgery groups (**Table 2**). Despite these household economic differences, all groups reported high participation in prenatal care visits with slight skewing towards earlier (first trimester) visits for the MBS group compared to both control groups. Parity was slightly less, on average, in the MBS group than in the control groups. While 95-96% of each group were “never” smokers, the MBS group reported a higher “never” alcohol use than either control groups (ASD 0.57 vs. 0.51 for both comparisons), although missingness was high for this variable.

The maternal comorbidity index indicated good balance (ASD <0.15) between the MBS and control groups with a slight skewing toward 2 or more comorbidities in the MBS group than the controls groups (**Table 2**). On the other hand, pre-gestational hypertension was more common in both the MBS and pre-surgical control groups (10.3%) than the pre-pregnancy control group (4.5%) (**Table 2**).

Like the baseline matching variables, differences in baseline demographic and maternal health variables were found between groups selected in the phase 2/3 matching process (**S4 Table**), possibly reflecting the greater skewing towards more extreme pre-surgical BMI’s in the MBS group vs. pre-surgery control groups.

## Discussion

The POSIT study is a retrospective cohort study designed to improve understanding of the effects of MBS on long-term child health outcomes. Leveraging multi-regional data from three integrated health care networks, we identified and characterized a cohort of 1,374 women with MBS treatment prior to pregnancy matched to 13,740 women with BMI similar to their pre-surgical BMI and 13,740 women with BMI similar to their pre-pregnancy BMI. Using this data resource, we will test the hypothesis that compared to offspring of pre-surgery control women, preconception MBS will lead to lower infant and childhood weight gain and blood pressure through age 6 years(**Fig 3**, **Table 3**). We also hypothesize that these infant and child parameters will more closely match the offspring of the pre-pregnancy control women. As a consequence of preconception weight loss after MBS, improvements in subsequent maternal GDM and hypertensive events, and improved gestational weight gain, will mediate beneficial outcomes on offspring obesity and chronic disease risk compared to pre-surgery controls (**Figure 3**, **Table 3**). In contrast, a potential mediator of increased offspring weight gain risk includes MBS-induced micronutrient malabsorption and worse maternal anemia as well as increased SGA frequency. The theorized mechanisms underlying these hypotheses include preconception weight loss by MBS leading to lower preconception maternal BMI, improved maternal metabolic control, with subsequent reduced exposure to epigenetic and programming pressures during conception and fetal development.

**Figure 3:**
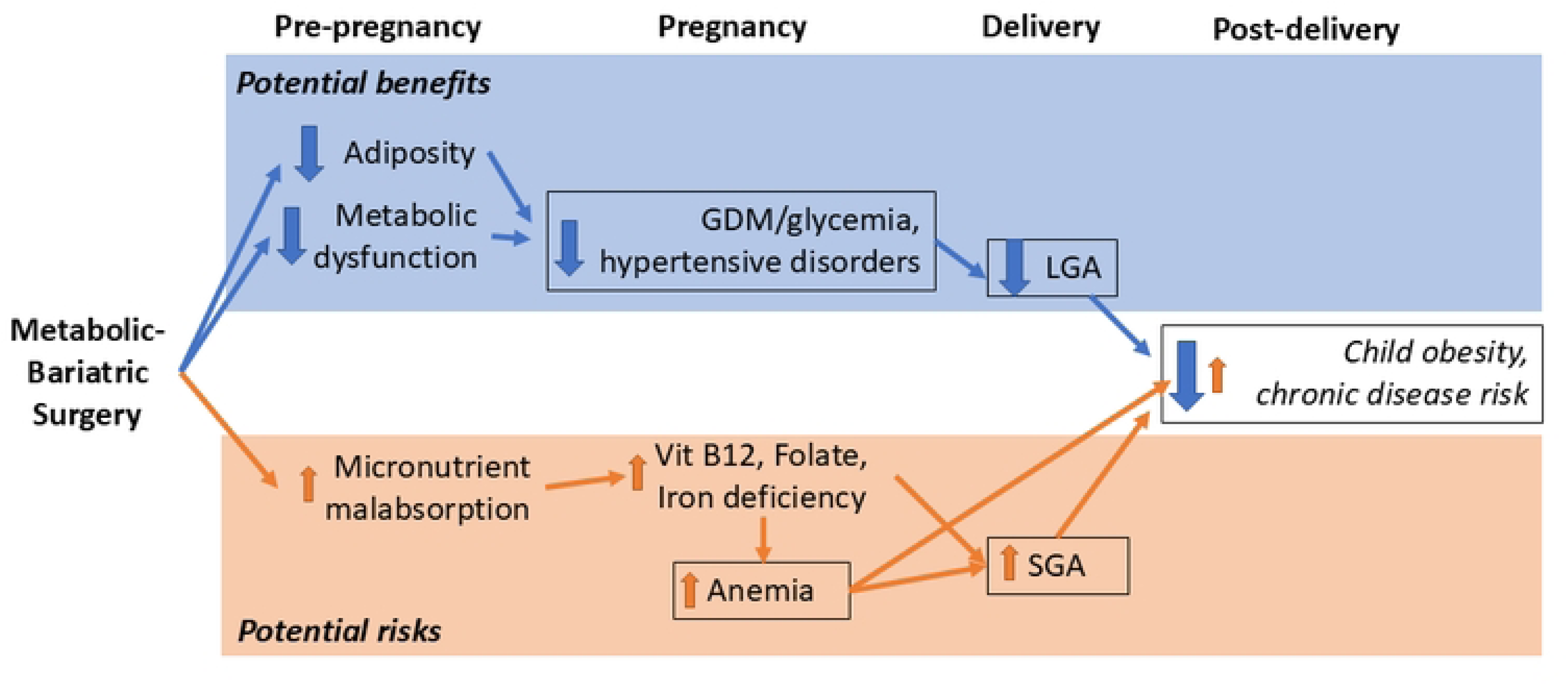
POSIT study hypotheses. While metabolic-bariatric surgery can increase risk for small for gestational age (SGA) infants, on balance, improvements in maternal risk for gestational diabetes (GDM) and hypertensive disorders after MBS are predicted to favor lower childhood risk for obesity and chronic diseases. LGA: large for gestational age.

**Table 3.**
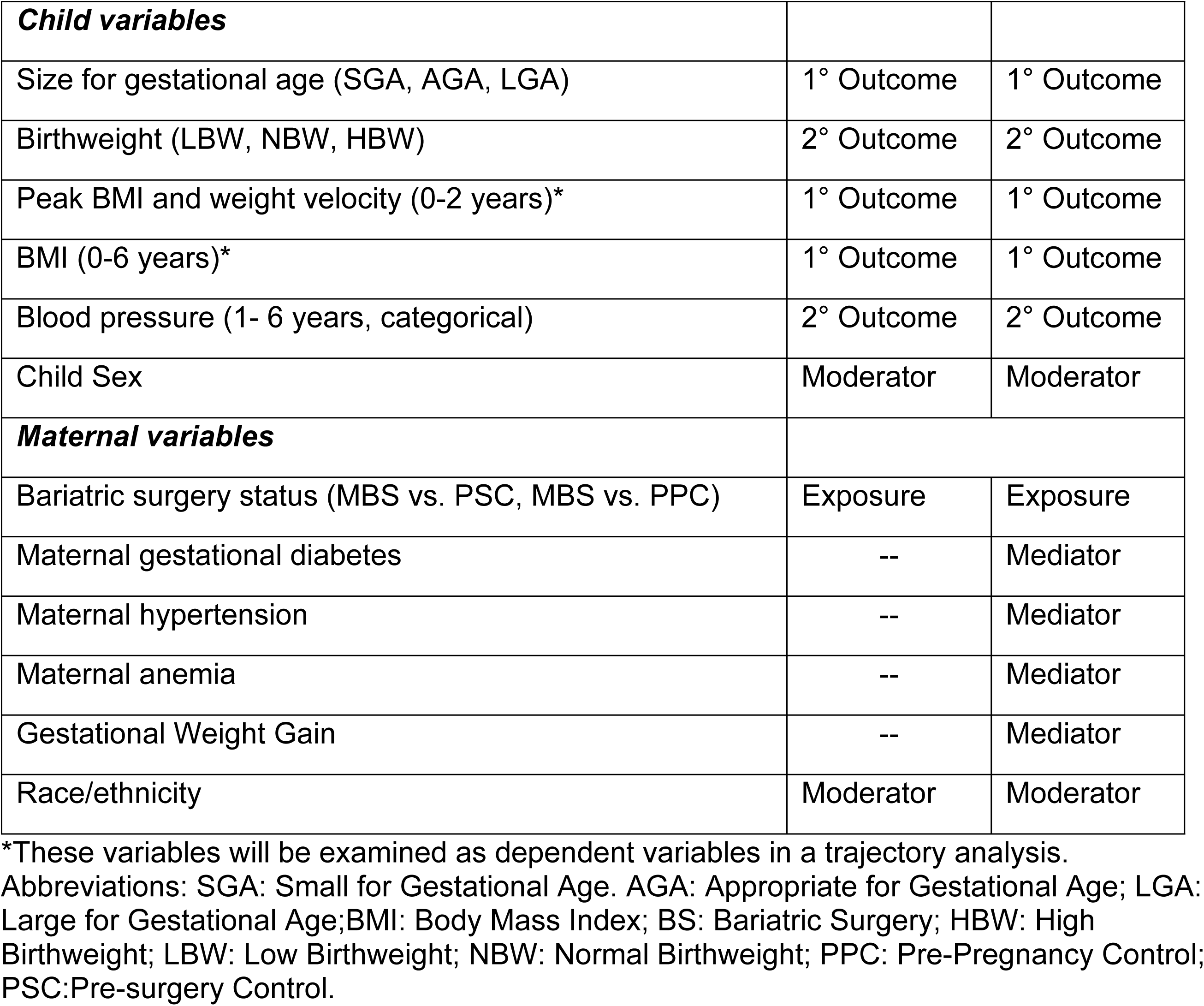
POSIT key outcome variables.

Only a handful of studies to date have explored the effect of surgically induced preconception weight loss and improved maternal metabolic control on long-term offspring health, including infant growth, propensity for obesity, and expression of weight-related comorbidities. While early studies were promising,[27] subsequent reports showed inconsistent benefits on childhood growth [28–30], and others did not demonstrate any benefit on offspring body weights.[31–33] Limitations in sample size and differences in methodologic approaches between studies likely account for these inconsistencies. None of these reports could investigate potentially important mediators of long-term benefit or harm, including changes after bariatric surgery in risk for maternal anemia, GDM, and hypertensive disorders of pregnancy. Findings of small for gestational age (SGA) infants in these studies were also inconsistent, even though this risk is higher in pregnancies post-bariatric surgery [24, 25] and is associated in the general literature with longer-term risks for childhood obesity and chronic diseases.[42, 43]

Given that a randomized, prospective study that assigns women-to-be to either MBS or placebo is not feasible, the POSIT study uses a matched design using data from three large health systems, providing longitudinal clinical data needed to fill these knowledge gaps. We considered alternative designs, including comparing children before and after MBS[44] or enrolling participants prospectively, but the matched cohort strategy offered the best opportunity to feasibly work with a large, prospective dataset adequately powered to test our hypothesis and that also included a racially and ethnically diverse group of women.

Using data from the POSIT cohort, we will compare infant and childhood weight and blood pressure outcomes in offspring of women who lost weight following metabolic-bariatric surgery versus one of two control groups with no previous bariatric surgery history. Planned secondary analyses of MBS treatment include three alternative exposure variables: time from surgery to conception (months between the date of surgery and estimated date of conception, classified as < 6 months, <12 months, and ≥ 12 months),[45, 46] amount of weight loss induced by the surgery (categorized by tertiles of percent weight loss between surgical dates and pregnancy), and bariatric surgery type (RYGB, SG). We plan to analyze three primary child anthropometry outcomes: size for gestational age, infant (from birth to 2 years age) weight gain (growth) trajectory, and BMI from birth to age 5. Birth weight (BW) classified as low (<2.5 kg; LBW), normal, and high (>4.5 kg; macrosomia) for full-term births will be examined as a secondary child outcome. Infant growth parameters will be examined as dependent variables, with peak BMI and BMI velocity as the primary parameter of interest. We will use the BMI of children between 2 and 5 years of age as an indicator of weight status at preschool-age and school entry. Finally, we will examine systolic (SBP) and diastolic (DBP) blood pressures obtained from clinical encounters when the child was 3 to 5 years of age.

In addition to examining child outcomes following a clinically important obesity treatment, POSIT study findings will also have implications for the broader field of the impacts of maternal obesity on childhood chronic disease prevalence and the extent to which they are independent of parental genetics.[47] One approach to disentangling the contributions of genetics from the *in-utero* environment on childhood obesity risk is to study the offspring of women with obesity who have achieved meaningful and sustained weight loss prior to conception. Because attaining clinically impactful preconception weight loss and maintenance through pharmacologic therapy or chronic caloric restriction is either difficult to achieve or could adversely impact fetal development, preconception weight loss by MBS for women with obesity offers a unique opportunity to study the effects of reducing maternal adiposity and associated metabolic dysfunction on subsequent child health. Studies of women who underwent MBS have shown that they have significantly lower risks of GDM and maternal hypertensive disorders (gestational hypertension, pre-eclampsia, and eclampsia) than women matched to their pre-surgery BMI, and similar to women matched to their pre-pregnancy BMI.[24, 25] These two control groups are important because the former allows estimation of the effects of weight loss on maternal-child outcomes, while the latter investigates if those effects are sufficient to achieve expected outcomes based on their new, lower weight.

Challenges to our protocol and analyses are that while we attempt to capture societal and environmental influences on childhood obesity risk, including social determinants, we will not be able to obtain specific maternal or child lifestyle information. Instead, we hope to mitigate cultural, population, and temporal influences on diet and activity by matching as closely as possible within the respective Kaiser Permanente site and to the year of pregnancy as possible, including adjusting for differences in baseline variables mentioned above and inclusion of a sensitivity analysis cohort. Finally, while examining growth parameters and blood pressure out to 5 years of age can provide a glimpse into future chronic disease risk,[48, 49] follow-up studies are needed to monitor health outcomes of these offspring cohorts into adolescence and beyond.

## Conclusion

Successful completion of the POSIT study will address limitations of previous studies leading to conflicting findings and restricted generalizability. It will also address concerns regarding the risks versus benefits of preconception bariatric surgery in women on their offspring’s long-term health. This is of major importance for clinicians counseling women-to-be with severe obesity, committees in charge of generating guidelines for the care of pregnant women, and policymakers establishing programmatic priorities to reverse the currently rising population trends of maternal obesity and accompanying maternal-fetal complications.

## Data Availability

Data are available upon reasonable request according to the following: Business Confidential or Proprietary Information, categorized as non-public information under KPSC’s Institutional Policy, are not made available in broadly accessible repositories. KPSC shall maintain the data in-house and agrees to share deidentified data according to the following policies and procedures that will provide data access to qualified researchers: researchers will need to submit a pre-application or preparatory to research data request that will need to be evaluated and approved by the investigators of the study and scientific directors at KPSC’s Department of Research & Evaluation, receive IRB approval, and have appropriate data use agreements set up.

## Acknowledgements

The authors would like to thank Dr. Matthew Daley for his valuable input during study conceptualization.

## Author contributions

JQP, DG, KKV, CEO, SQ, JMS, and JBH conceptualized the study design and approach. CPW, SQ, PK, and TMI were responsible for project administration and data curation. SQ and JMS performed formal statistical analysis. JQP and JBH performed the original draft preparation and all authors contributed to manuscript review and editing.

## Supporting information

**S1 Table. Diagnosis and procedure codes used to identify metabolic-bariatric surgery dates.**

**S2 Table. Base population with a metabolic-bariatric surgery code and known surgery date by study site.**

**S3 Table. Baseline matching characteristics of POSIT cohorts included for sensitivity analysis.** Comparisons include the treatment group who underwent pre-conception metabolic-bariatric surgery, a pre-surgery control group matched to pre-MBS BMI, and a pre-pregnancy control group matched to BMI at the time of pregnancy.

**S4 Table. Baseline demographic and maternal health characteristics of POSIT cohorts included for sensitivity analysis.** Comparisons include the treatment group who underwent pre-conception metabolic-bariatric surgery, a pre-surgery control group matched to pre-MBS BMI, and a pre-pregnancy control group matched to BMI at the time of pregnancy.

